# The genomic and epidemiological virulence patterns of *Salmonella enterica* serovars in the United States

**DOI:** 10.1101/2022.12.13.22283417

**Authors:** Gavin J. Fenske, Jane G. Pouzou, Régis Pouillot, Daniel D. Taylor, Solenne Costard, Francisco J. Zagmutt

**Affiliations:** EpiX Analytics, Fort Collins, Colorado, United States of America

## Abstract

The serovars of *Salmonella enterica* display dramatic differences in pathogenesis and host preferences. We developed a process (patent pending) for grouping *Salmonella* isolates and serovars by their public health risk to provide better S*almonella* control targets along the food chain. We collated a curated set of 12,337 *S. enterica* isolate genomes from human, beef, and bovine sources in the US. After annotating a virulence gene catalog for each isolate, we used unsupervised random forest methods to estimate the proximity (similarity) between isolates based upon the genomic presentation of putative virulence traits We then grouped isolates (virulence clusters) using hierarchical clustering (Ward’s method), used non-parametric bootstrapping to assess cluster stability, and externally validated the virulence clusters against epidemiological virulence measures from FoodNet, the National Outbreak Reporting System (NORS), and US federal sampling of beef products. We identified five stable virulence clusters of *S. enterica* serovars. Cluster 1 (higher virulence) serovars yielded an annual incidence rate of domestically acquired sporadic cases roughly one and a half times higher than the other four clusters combined (Clusters 2-5, lower virulence). Compared to other clusters, cluster 1 also had a higher proportion of infections leading to hospitalization and was implicated in more foodborne and beef-associated outbreaks, despite being isolated at a similar frequency from beef products as other clusters. We also identified subpopulations within 11 serovars. Remarkably, we found *S.* Infantis and *S.* Typhimurium subpopulations that significantly differed in genome length and clinical case presentation. Further, we found that the presence of the pESI plasmid accounted for the genome length differences between the *S.* Infantis subpopulations. Our results demonstrate that *S. enterica* strains with the highest incidence of human infections share a common virulence repertoire. This work could be used in combination with foodborne surveillance information to best target serovars of public health concern.

## Introduction

Members of *Salmonella enterica* subspecies *enterica* are some of the most ubiquitous agents implicated in foodborne human illnesses. Despite being constituents of the same subspecies, members of *S. enterica* are not only commonly isolated from livestock but also amphibians [1] and wild birds [2]. The wide host range for *S. enterica* makes control of the pathogen exceedingly difficult due to the large number of potential reservoirs. Historically, strains of *S. enterica* have been grouped into units termed serovars based upon serological antigen presentation. While an initial list presented 44 *S. enterica* serovars in 1934 [3], today’s descriptions include over 2,500 serovars of *S. enterica* [4]. Nonetheless, in the US only 20 serovars accounted for 69.2% of human *S. enterica* isolates collected in 2016 by the US Centers for Disease Control and Prevention’s (CDC) Laboratory-based Enteric Disease Surveillance (LEDS) program [5]. Furthermore, nearly 10% of *S. enterica*’s serovars may be polyphyletic or paraphyletic [6].

To establish infections in disparate hosts, *S. enterica* manipulates common immune functions of higher vertebrates. Indeed, the classic gastroenteritis associated with *S. enterica* infections is the result of the pathogen affecting the host’s innate immune system to generate inflammation, subsequently producing unique metabolic niches for *S. enterica* while killing its competitors for reduced substrates in the hindgut [7-9]. Such remarkable expropriation of the hosts immune functions is achieved by virulence genes (virulence factors), many of which are contained within chromosomal elements termed Salmonella Pathogenicity Islands (SPI) [10]. Genes contained within SPI aid in host cell invasion, and subsequent survival and dissemination within and between Eukaryotic host cells [11,12]. However, serovars display differences in pathogenesis and host-preferences. For example, the human-restricted serovar *S. enterica* ser. Typhi (*S.* Typhi), the etiological agent of typhoid fever, does not typically cause submucosal inflammation and resultant diarrhea in infected patients as with classical salmonellosis, but instead elicits a systemic enteric fever characterized by initial immune evasion [13,14]. *S.* Dublin, a bovine adapted serovar, commonly generates systemic infections in humans and is isolated from blood samples in 61% of human clinical infections as compared to an average of 5% for other *S. enterica* serovars in the US [15]. The general pathogenesis of *S. enterica* is not fully elucidated, and the virulence potential for individual serovars is poorly understood. Furthermore, most studies have focused upon *S.* Typhimurium as a model organism for all *S. enterica* virulence, [8,16-21] which could obfuscate differences between serovars.

Despite the tremendous virulence diversity within *S. enterica*, microbial criteria from the US Food Safety and Inspection Services (FSIS) on important sources of *S. enterica* such as beef and poultry meats target all serovars equally, based on prevalence. Further, traditional surveillance methods can take considerable time to identify emerging serovars of public health concern, thereby delaying food safety intervention implementation [22]. Understanding virulence differences between serovars and identifying emerging virulent serovars in a timely manner can inform more focused risk management strategies targeting serovars with an inordinate impact on public health while reducing food waste due to recalls.

Previous studies have used genomics to identify serovar groups of public health concern. Karanth et al. analyzed a limited number of genomes and serovars originating from humans, poultry, and swine to characterize virulent serovars [23]. This analysis had the benefit of using the entire genome of Salmonella to group isolates by disease presentation; however, the computational resources required prevent its application to a large number of isolates. In another study, researchers used single nucleotide polymorphism (SNP) clusters and *S*. Saintpaul as a model to identify virulent isolates [24]. Although using high-resolution genomic methods identified SNP clusters associated with a high proportion of human clinical isolates, *S*. Saintpaul may not be the best serovar model due to its polyphyletic nature [25]. The objective of this study was to develop a computationally efficient genomic approach to group *Salmonella* serovars by virulence biomarkers in isolates from humans, beef, and bovine animals and define the human health risk of the resulting clusters using epidemiological data. We chose beef as a model foodstuff since US federal monitoring of *Salmonella* in beef is well-established, nationally representative, and beef remains an important vehicle for *S. enterica*. Beef production in the US is more decentralized than poultry and pork production [26] and we hypothesize that this decentralization may present unique genomic populations arising from geographic separation as previously observed in *S. enterica* serovars [27,28]. Furthermore, *S. enterica* in beef products is understudied compared to other vehicles such as eggs, poultry, and pork meat.

## Materials and methods

We developed an information pipeline (patent pending) using virulence factors as markers and epidemiological data as validation to group serovars by their risk to human health. After compiling a curated set of *S. enterica* genomes (n=12,337) from human, bovine, and beef sources, we applied an unsupervised random forest and hierarchical clustering approach to group isolates based upon genomic virulence trait presentation and validated the groups against epidemiological measures including clinical presentation from sequenced isolates collected by the FoodNet active surveillance network (29).

### Contig assembly selection and quality criteria

We compiled *S. enterica* assemblies from bovine-associated isolates from three primary sources: 1. BioProject PRJNA242847 (FSIS HACCP samples, accessed 7/13/2021), 2. BioProject PRJNA292666 (FSIS NARMS isolates, accessed 7/13/2021), and 3. BioProject PRJNA292661 (FDA NARMS isolates, accessed 8/25/2021). We collected isolates from sources specified as bovine-associated or beef origin from the metadata for the above BioProjects.

We retrieved *S. enterica* isolates from human clinical cases from BioProject PRJNA230403 (CDC PulseNet, accessed 9/13/2021) and identified sporadic, domestically acquired, non-attributed *S.* enterica isolates from the FoodNet active surveillance network [29]. We did not include outbreak cases from FoodNet since they are not attributed to a particular source in that dataset. Instead, we used the National Outbreak Reporting System (NORS) dataset, a passive system for reporting enteric disease outbreaks in the US, to identify additional human isolates that originated from beef-attributed outbreaks. We initially defined beef attribution based on the Interagency Food Safety Analytics Collaboration (IFSAC) classification. As IFSAC-defined beef-associated salmonellosis outbreaks for which clinical isolates were sequenced are limited, we widened the definition of potentially beef-associated illness to include outbreaks which listed beef as an identified contaminated ingredient. We based this inclusion on whether the list of foods or ingredients per outbreak included beef, even if other possible ingredients could not be ruled out to definitively assign an IFSAC classification. The following text strings were used to identify beef-associated outbreaks: "beef", "burger", "steak", "carne", "kitfo", "ox tongue", "short-rib", "prime rib", "barbacoa". If the IFSAC classification attributed an outbreak to other foods, we did not designate it as a beef-associated outbreak.

We removed isolates from the data set if: 1) No pre-computed assembly was available on NCBI, 2) SKESA v. 2.2 was not used to construct the assembly, 3) The number of contigs representing the assembly was greater than 300, and 4) The contig n50 was less than 25,000 bp. After initial parsing for isolation sources and assembly quality, we included serovars with 50 or more isolates in the analysis. In total, the final analysis set includes 12,337 assemblies and represents 37 serovars.

### In silico serovar prediction

We used *Salmonella in silico* Typing Resource (SISTR) [30] with default options to assign putative serovars to each assembly. 1,077 assemblies failed the quality control step within the SISTR software with the same error message “FAIL: Wzx/Wzy genes missing…”, but all 330 / 330 genes for the core genome multilocus sequence typing (cgMLST) scheme used within the software were present within these assemblies. We retained assemblies failing QC with the aforementioned error message and that contained all 330 cgMLST loci for the analysis while excluding any assemblies which failed the quality control step and did not have all cgMLST genes. S1 table provides the SISTR serotyping results for each assembly.

### Virulence gene annotation

We collated a custom database of putative virulence factors associated with *Salmonella*, *Escherichia*, *Shigella*, and *Yersinia* from the virulence factor database (VFDB) (accessed 9/13/2021) [31] and putative virulence factors associated with *Salmonella*, *Escherichia*, and *Shigella* from PATRIC (accessed 9/13/2021) [32]. Next, we combined amino acid sequences of the open reading frames (ORF) with a reference proteome of *Salmonella* Typhimurium LT2 (https://www.uniprot.org/proteomes/UP000001014) and made the database non-redundant by clustering the open reading frames at 0.90 global identity using cd-hit [33]. We passed the resultant database to Prokka [34] using the “--proteins” option to specify the database as the primary annotation database in the software pipeline. We then parsed gene annotations from the VFDB and PATRIC non-redundant database from the resultant Prokka annotation tables. Additionally, to ensure consistent ORF predictions between assemblies, we trained a model using Prodigal [35] on the chromosome of the reference Salmonella Typhimurium LT2 assembly ASM694v2 (https://www.ncbi.nlm.nih.gov/assembly/GCF_000006945.2/) and passed the training file to Prokka using the command “--prodigaltf”.

### pESI plasmid identification

The pESI plasmid is an emerging concern in some *S. enterica* serovars, namely *S*. Infantis [36]. To confirm the presence of this plasmid, we complied an additional nucleotide database of 13 marker genes previously used to identify pESI plasmids in *S. enterica* contig assemblies from two sources [36,37]. We then conducted a nucleotide BLAST search against the database using the software Abricate [38] and defined positive hits as a percent identify of ≥ 95% and percent coverage of ≥ 95%. We presumed that an isolate contained the pESI plasmid if the contig assembly was positive for the pESI specific *repA* gene and contained at least five additional marker genes.

### Random forest model construction

We used virulence factor gene annotations from the resultant Prokka outputs and constructed a count matrix of virulence genes (assemblies x virulence factors). We excluded putative virulence factors present in more than 95% of assemblies or which were not present in at least 10 (0.08%) assemblies. To generate row similarity (isolate relatedness), we fit an unsupervised random forest to the count matrix of virulence factors using the randomForest [39] package in R[40]. The random forest model contained 50,000 trees and used 60 features (columns, virulence factors) at every split.

### Grouping isolates and assessing cluster stability

We converted the row-wise proximity matrix from the random forest model to a distance matrix (1 – similarity) and subjected it to agglomerative clustering using Ward’s method [41]. We conducted two analyses: 1) k = 5, used to group serovars by putative virulence factor patterns and 2) k = 37, using the same number of clusters as serotypes in the data to identify possible subpopulations within serovars with different virulence factor catalogues. We defined cluster stability for both analyses as a Jaccard similarity of ≥ 0.75 [42] for 10,000 non-parametric bootstrap samples. For the main serovar clustering analysis, we choose k=5 as it was the highest value for which all clusters were stable. The k=37 version of the analysis tested whether distribution of virulence factor combinations is more similar within each serotype than between serotypes and, therefore, whether serotype is a reasonable representation of these virulence differences. We defined serovar subpopulations as serovars with at least two of these populations annotated into different clusters (k = 37 clustering) and with at least two of the populations representing greater than or equal to 0.20 of the total serovar population. Clustering and bootstrapping were performed using the clusterboot function from the fpc [43] package in R.

### Epidemiological indicators

We estimated epidemiological indicators for both virulence clusters and serovars using sporadic and domestically acquired cases from FoodNet (2016 – 2019). We excluded outbreak-associated cases to decrease bias due to outbreak size and removed travel-related cases to exclude foodstuffs from regions that may have different *S. enterica* population structures than the US and are not good indicators of US consumer exposure to *S. enterica* in food. We calculated the proportion of positive samples for *Salmonella*, individual serovars, and each cluster in beef products from FSIS regulatory testing programs: MT43 (raw ground beef), MT60 (manufacturing trimmings) and MT64 (Components other than trim) for 2016 – 2019. We modeled all proportions using *Beta*(s+0.5, n-s+0.5) (eqn. 1), with a Jeffery’s prior (*Beta*(0.5, 0.5) as a Bayesian conjugate to the Binomial distribution[44]. Table 1 lists parameters *s* and *n* used to model these proportions. We used 1M Monte Carlo simulations to estimate and compare posterior distributions using numerical integration, with a 99% confidence level for statistical significance.

**Table 1.**
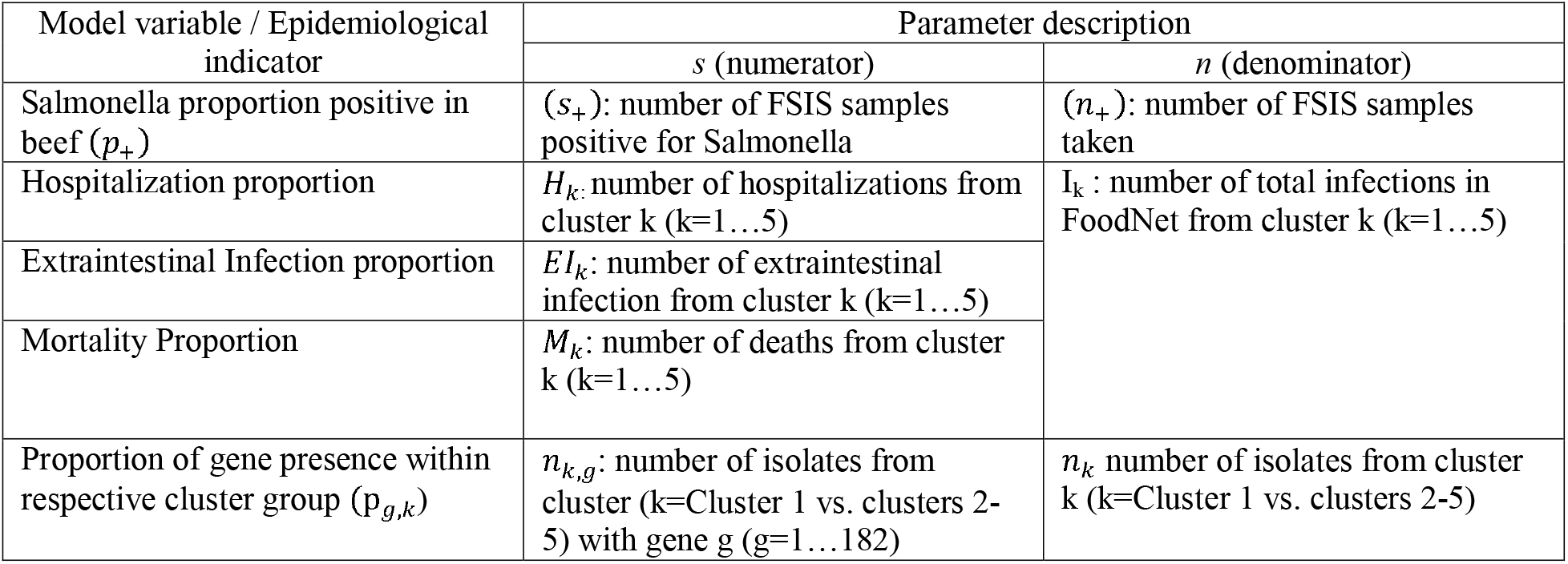
Description of parameters used for calculation of epidemiological indicator estimations using Equation 1.

#### Incidence of domestically acquired sporadic cases

We modeled the incidence of domestically acquired sporadic cases per 100k people per year (*λ_ij_*) for serovar *j* in FoodNet state *i* using the Bayesian conjugate for a Poisson rate with Jeffrey’s prior *Gamma*(0.5, 0.00001)(44), hence Gamma(α*_i_* + 0.5, β*_i_*t + 0.00001)(eqn. 2), where α_ij_ is the serovar case totals per state and β*_i_* is the FoodNet catchment area population for *t*=4 years. Sporadic cases are defined as illnesses which were not linked to a known outbreak. The catchment area of FoodNet is not evenly distributed between the 10 participating states, so the population-weighted mean serovar incidence (*λ_j_*) for the study period was 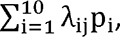 where *λ_ij_* is the FoodNet state-specific mean serovar incidence and p*_i_* is the state catchment proportion of total FoodNet population. Virulence cluster population-weighted average incidence, *λ_k_*, is the sum of the clusters’ constituent serovars incidence rates for the study period 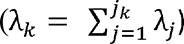.

#### Serovar proportion positive in beef

We determined the proportion of *Salmonella* positives (p_+_) following eqn. 1, with *s_+_* number of samples positive for *Salmonella* and *n_+_* total number of samples from FSIS testing. We estimated the proportion of serovar *j* isolated from beef products (x*_j_*) with a Dirichlet distribution, (*Dir*(a_j_) eqn. 3), where a_j_ is the number of FSIS isolates from serovar *j*. We excluded serovars without any positive isolates in FSIS testing and retained all serovars from the testing program (including those not included in the analysis set).

Serovar proportion positive (p*_j_+__*) was taken as the product of total *Salmonella* proportion positive (p_+_) and the serovar proportion of the total *Salmonella* population from eqn. 3 (x*_j_*). Finally, we derived the cluster proportion positive (p*_k_+__*) as the summation of the cluster’s constituent serovars.

#### Hospitalization, Extraintestinal Infection, and Mortality Proportions

We determined the proportion of infections with a certain outcome (i.e., hospitalization, extraintestinal infections, and mortality) for each cluster *k* (k=1…5) and for cluster 1 vs the combinations of others. We defined extraintestinal infections as having "URINE", "BLOOD", "ORTHO", "ABSCESS", "OTHER STERILE SITE" and "CSF" isolation sources. We modeled all the proportions using eqn. 1, with parameters described in Table 1.

### Differential gene carriage

To identify virulence factors differential between clusters, we trained a supervised random forest model (ntree = 5,000, features to try at split = 13) to classify isolates into two groups: cluster 1 (higher virulence) and clusters 2-5 (lower virulence). We extracted variable importance from the random forest model and defined factor importance using the mean decrease of Gini impurity. As with other proportions, we used eqn. 1 to model the proportion of factor presence (p_*g,k*_) within respective cluster group (1 vs. 2-5) for each of the virulence factors used in the random forest. The relative frequency (RF) of a given factor was the resultant ratio of proportions of factor presence (RF = p_*g*_1__ /p_*g*_(2-5)__). To ease interpretation of differential genes, we categorized them into five broad virulence mechanism categories (Adhesion, Motility/Invasion, Survival/Host Persistence, Toxin and Virulence Factor Secretion) (Table S3) based on their virulence descriptions listed in the VFDB [31] and PATRIC [32] databases.

### Code and data availability

Aggregated data and code used to generate figures for this study are available in our online repository: (link will be provided upon publication, Files shared with reviewers). FoodNet data is used with permission from the Centers for Disease Control and Prevention and although raw data may not be shared, code written from aggregated inputs is provided in our online repository.

## Results

### Genome assemblies analyzed

The Pathogen Detection Network hosted by NCBI contains over 400,000 sequenced *Salmonella* isolates from various sources and contributors. From these, we extracted 53,849 isolates from specific sampling programs. We further reduced to a final analysis set of 12,337 *S. enterica* assemblies comprised of 37 serovars representing human clinical cases in the US and bovine and beef associated isolates, (Fig 1). Approximately 55% (6,751) assemblies are from US human clinical infections with the remaining 45% (5,586) representing isolates from bovine animals and beef products. The metadata for the genomes analyzed is provided in S1 Table.

**Fig 1.**
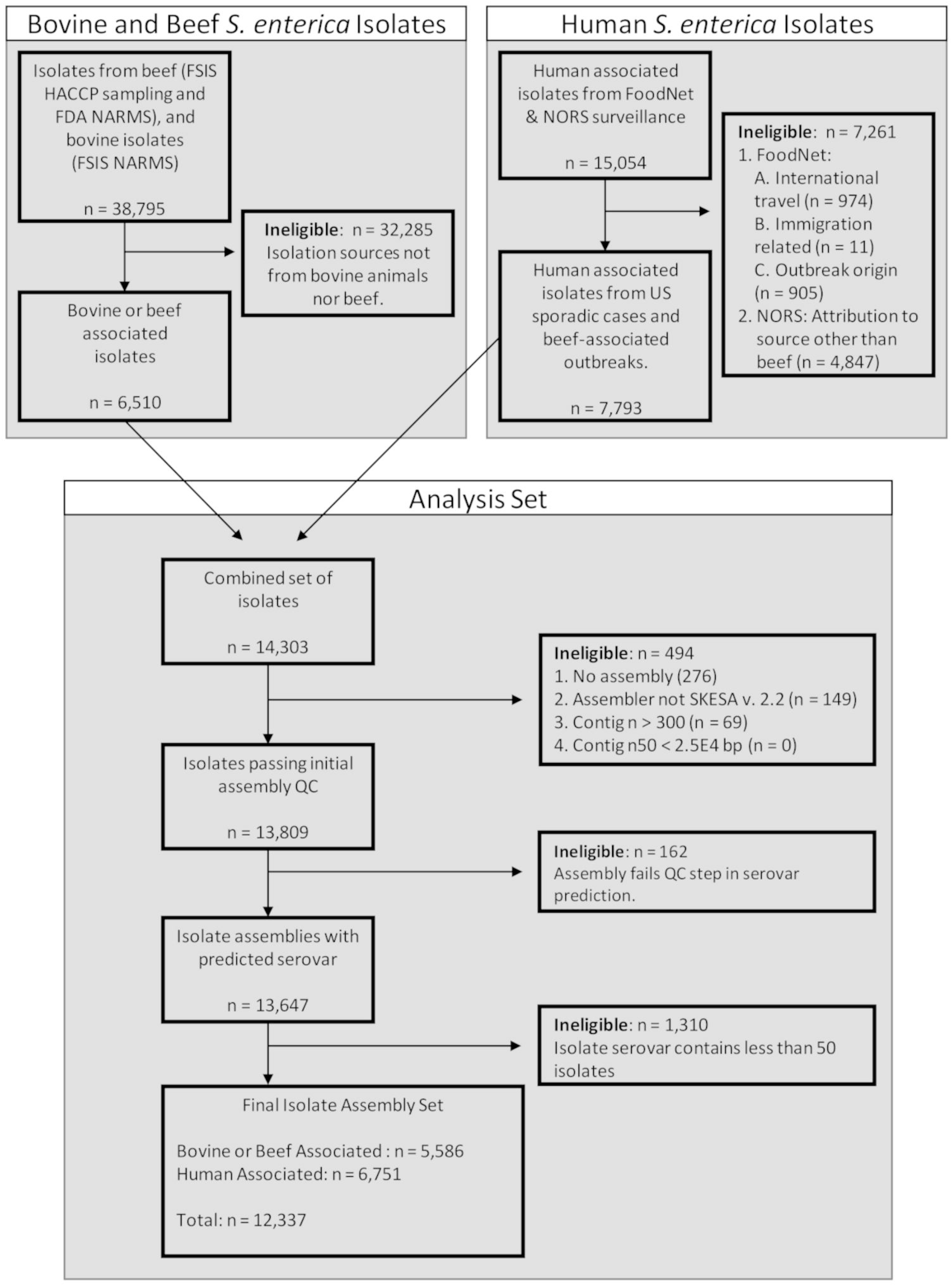
Analysis set of genomes. Description of the *S. enterica* genome assemblies considered, and exclusion and inclusion criteria applied to generate the analysis set.

### Clustering serovars using isolate virulence gene catalogues

To establish clusters of serovars, we identified virulence factors genes from each assembly and compiled them into a count matrix, trained an unsupervised random forest model to approximate similarity between isolate virulence factor catalogues (Fig 2), and subjected the resultant isolate similarity matrix to agglomerative clustering to identify clusters with subsequent non-parametric bootstrapping to validate cluster stability.

**Fig 2.**
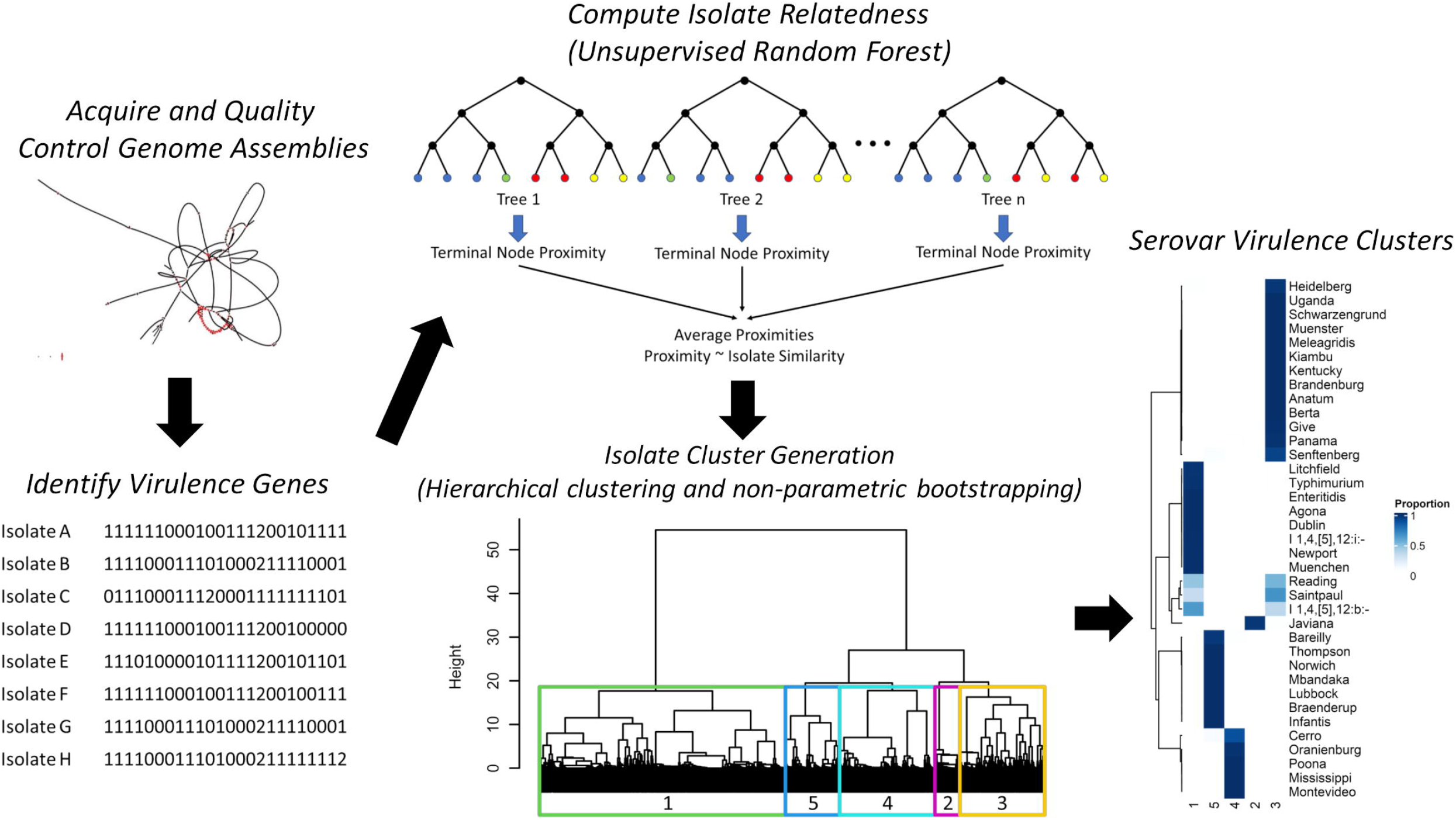
Conceptual model of virulence cluster development. First, we downloaded contig assemblies and quality controlled for fragmentation followed by the identification of virulence factors. We then fit an unsupervised random forest model to the isolate level virulence factors catalogues to approximate relatedness. We converted the resultant similarity matrix to a distance matrix (1 – similarity) and clustered using Ward’s method. We identified five stable clusters and validated using non-parametric bootstrapping.

We identified five stable clusters of *S. enterica* isolates (Fig 3A), with the majority of serovar isolates residing within the same clusters (mean within serovar cluster proportion = 0.96). (Fig 3B). However, *S.* Reading (cluster 1: n = 28 (0.47), cluster 3: n = 32 (0.53)), *S.* Saintpaul (cluster 3: n = 134 (0.66), cluster 1: n = 68 (0.34)), and *S.* I 1,4,[5],12:b:- (cluster 1: n = 53 (0.66), cluster 3: n = 30 (0.34)) had at least 33% of total serovar isolates in two different virulence clusters. The five virulence clusters are of uneven size (Fig 3C) with cluster 1 containing almost 10 times more assemblies than cluster 2. We attempted to decrease the size of cluster 1 by introducing a sixth cluster. However, the sixth cluster was unstable (bootstrap Jaccard similarity = 0.515) and cluster 4 was split, not cluster 1, indicating that the variance (Ward’s method used to cluster) within cluster 1 is less than that of cluster 4 despite its much larger size (S1 Fig). Interestingly, cluster 2 is comprised of only *S.* Javiana and the cluster homogeny of *S.* Javiana was preserved with the addition of the sixth cluster (S1 Fig). Serovar cluster designations are provided in Table S2.

**Fig 3.**
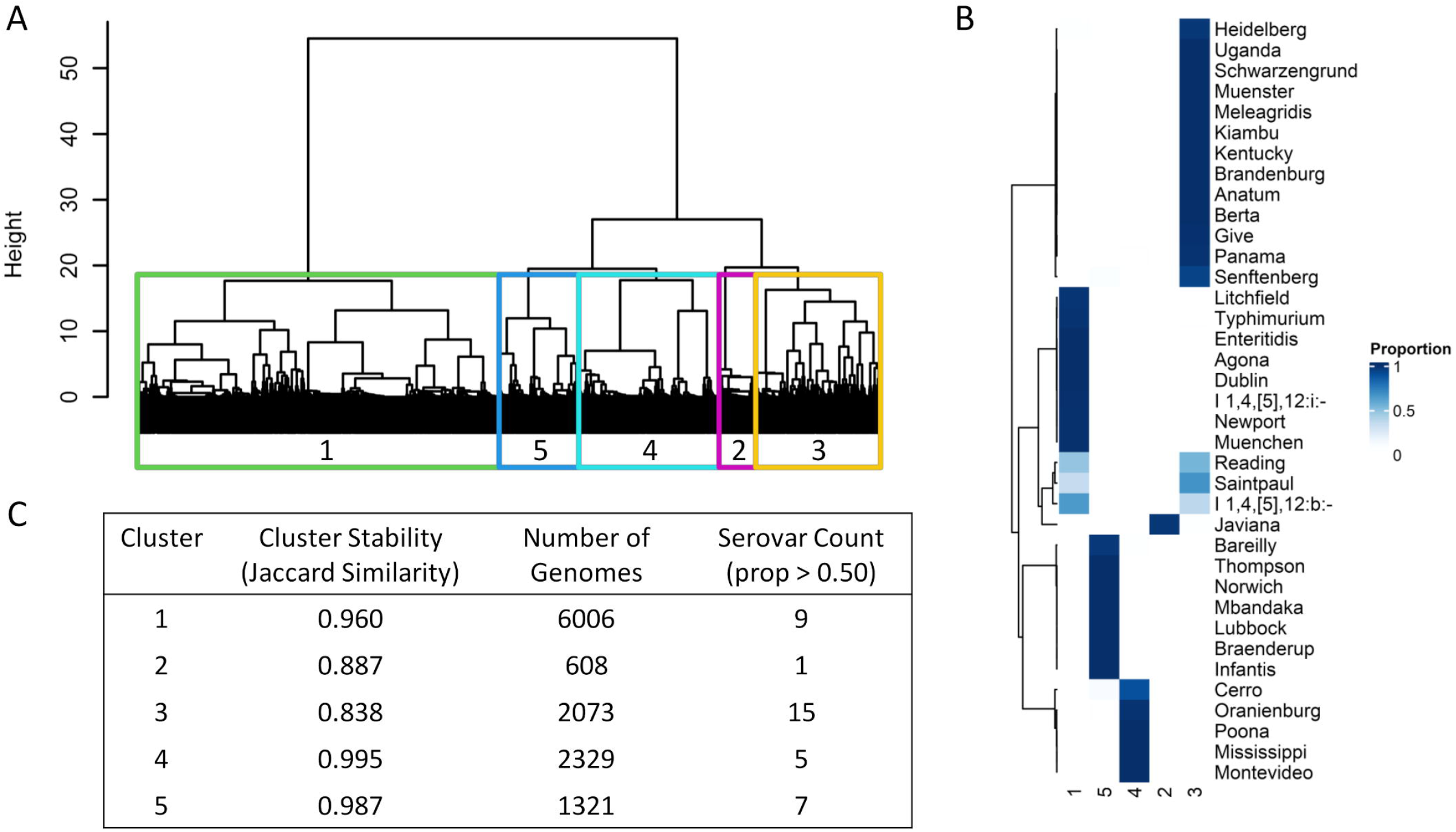
Description of the five virulence clusters. (A) Dendrogram depicting the hierarchical relationship between 12,337 *S. enterica* genome assemblies based upon virulence factor gene carriage with the five virulence clusters superimposed on top. (B) Heatmap of serovar proportion within each of the five respective virulence clusters. Rows are clustered using Ward’s method. (C) Characteristics of the five virulence clusters: cluster stability - Jaccard similarity of 10,000 non-parametric bootstraps, Number of Genomes - depicting the number of S. enterica genomes constituent in each cluster, and number of serovars (within cluster serovar proportion > 0.5) in each cluster.

### General epidemiological characteristics of virulence clusters

To investigate if the genomic virulence clusters correspond to clinical case presentation, we computed basic epidemiological characteristics per cluster for 2016-2019 as proxies for virulence phenotypes: proportion positive in beef products, number of outbreaks, incidence of domestically acquired sporadic cases per 100k people per year, hospitalization proportion given infection, extraintestinal infection proportion given infection, and mortality proportion given infection. We computed the results by virulence cluster (S4 Table). and by serovar. Not every *S. enterica* captured during surveillance programs in the US is subjected to sequencing, therefore we attributed cases from a given serovar to the cluster to which the highest proportion of serovar isolate was assigned (e.g., 98.5% of *S.* Typhimurium isolates were resident in cluster 1, therefore all cases of *S.* Typhimurium in the datasets were allocated to cluster 1). Cluster 1 serovars have the highest incidence rate of domestically-acquired sporadic cases (5.9 cases per 100k population per year, 99% CrI: 5.77 – 6.06) (Fig 4A), approximately 1.5x higher than that of clusters 2-5 combined during 2016 – 2019 (incidence rate ratio: 1.5, 99% CrI: 1.44 – 1.55). Moreover, infections from serovars in cluster 1 had a higher proportion of hospitalizations than serovars in cluster 2 (relative frequency (RF): 1.10, 99% CrI: 1.002 – 1.200), cluster 4 (RF: 1.15, 99% CrI: 1.029 – 1.296), and cluster 5 (RF: 1.17, 99% CrI: 1.058 – 1.288) (Fig 4B).The cluster 1 proportion positive in beef products was less than half of clusters 3-5 (proportion positive ratio: 0.44, 99% CrI: 0.366 – 0.528) (Fig 4C). However, cluster 1 serovars were implicated in the highest proportion of total foodborne outbreaks and beef associated outbreaks in the US from 2016 – 2019 (Fig 4D), generating approximately 2.5x more beef associated outbreaks (20 vs. 8) than clusters 3-5 combined (There were no cluster 2 (i.e., *S.* Javiana) isolates found in beef sampling or in beef associated outbreaks). Additionally, higher virulence serovars were involved in approximately 1.47x more foodborne outbreaks than clusters 2-5 combined from 2016 - 2019 (285 vs. 194).

**Fig 4.**
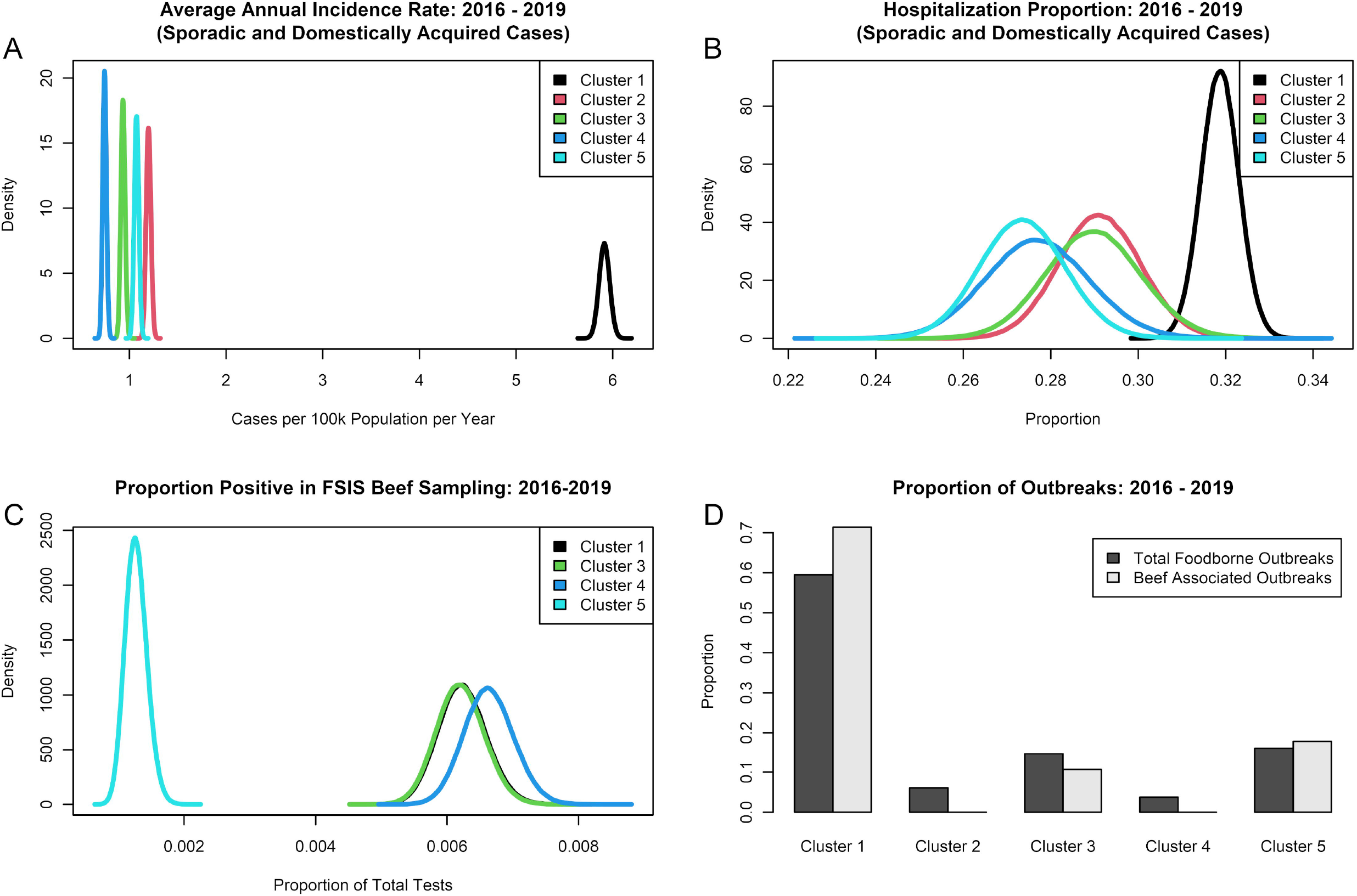
Epidemiological indicators of the five virulence clusters for the study period 2016-2019. (A) Incidence of domestically acquired sporadic cases per 100k population per year by virulence cluster. (B) Proportion of clinical infections resulting in hospitalization by virulence cluster. (C) Proportion positive estimates in FSIS testing of US beef products. No isolates from cluster 2 (comprised solely of *S.* Javiana) were retrieved from 2016 – 2019. (D) Proportion of total US foodborne and beef-associated outbreaks attributed to serovars in the analysis set.

### Differential carriage of virulence factors between clusters

In addition to the clear difference in epidemiological characteristics, a clear bifurcation exists between cluster 1 and clusters 2-5 (Fig 3A). We sought to identify virulence mechanism categories driving this differentiation by visually exploring the abundance (number of isolates carrying at least one copy of the virulence gene), frequency (RF), and clustering influence (mean Gini impurity) of the 182 virulence genes used in our analysis (Fig 5). The top two quadrants of the figure include the virulence factors that provided the most separation between clusters (highest mean Gini impurity), and the virulence factors on the two right quadrants were more common in the higher virulence group than in the lower virulence group (higher RF). Consequently, the upper-right quadrant includes factors that best distinguish the higher virulence cluster and were most frequent in the higher virulence group. Interestingly, factors involved in adhesion were the most important differentiators between clusters, while also being present in both virulence groups. Expectedly, more abundant genes generally provided higher differentiation, as some genes were very rare (e.g., 11 isolates contained the ompF gene), while others, such as the ratB gene, were found in up to 95% of isolates, as dictated by our gene exclusion criteria. Besides adhesion, other influential genes were involved in survival/host persistence, and motility/invasion, whereas genes manifesting toxin production provided less differentiation between virulence groups (Fig 5). The full list of virulence factors s considered in the analysis and gene metadata are provided in Table S3, while genes present in each isolate (n=68) are presented in Table S5.

**Fig 5.**
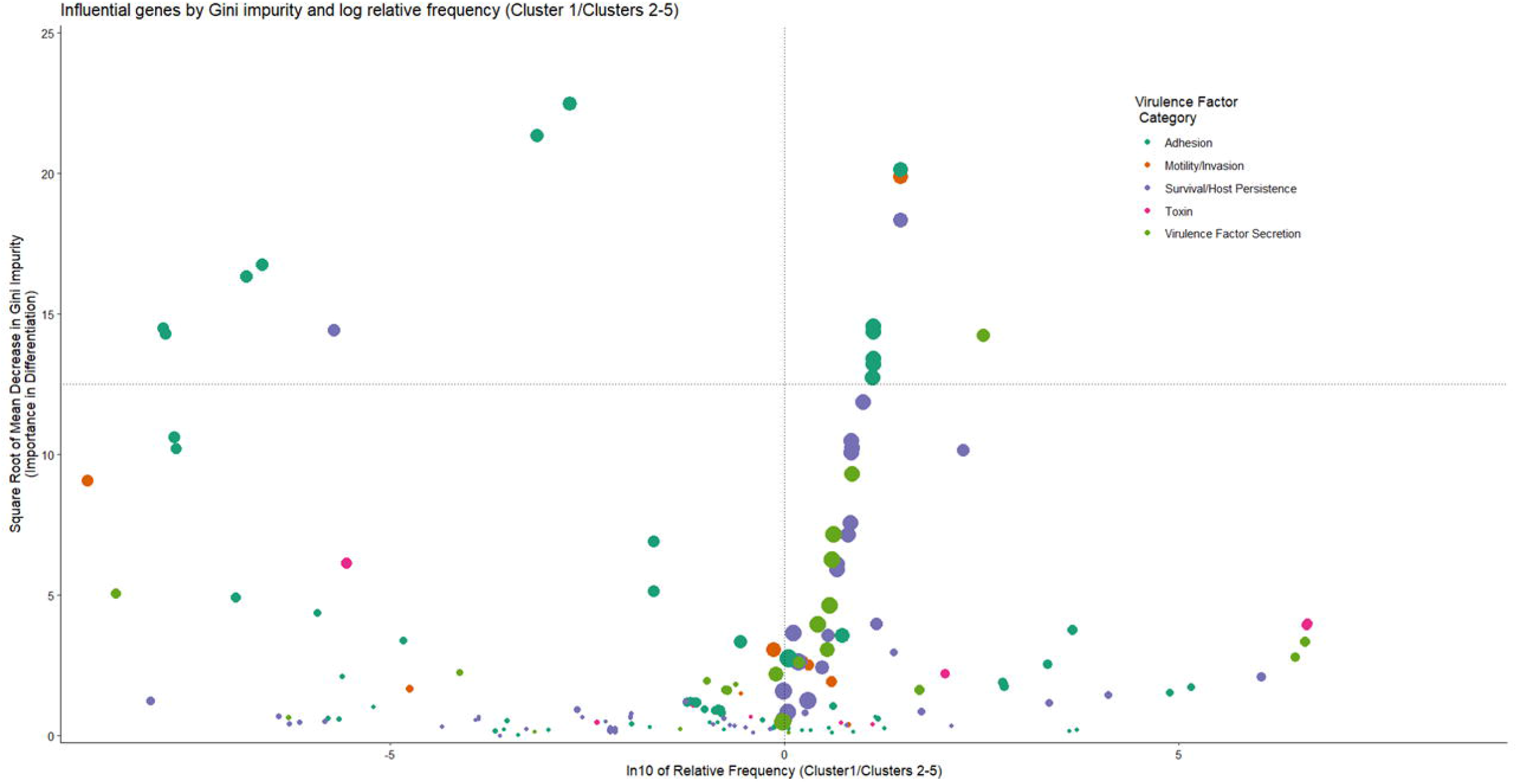
Abundance, relative frequency, and influence of 182 virulence factors used to classify 12,337 *S*. enterica genomes from human, beef, and bovine sources into two virulence groups. Points represent the square root mean decrease Gini impurity and natural log relative frequency (Cluster 1/Cluster 2-5) of each virulence factor, with diameter proportional to the number of isolates (min: 11, max: 11,653) carrying at least one copy of the virulence gene. Color designates virulence mechanism categories, as derived from the VFDB [31] and PATRIC [32] databases. Vertical dashed line represents equal frequency of virulence factors between the clusters, while points to the right of this line represent factors more frequently found in cluster 1. Points above the horizontal dashed line (square root of the mean decrease Gini impurity=12.5) represent virulence factors that were more influential differentiators in classifying isolates.

### Within serovar virulence subpopulations

Horizontal gene transfer molds virulence gene carriage, especially within SPI [45,46]. We hypothesized that horizontal gene transfer may lead to virulence subpopulations that could be identified using random forest methods otherwise missed in more traditional alignment-based phylogeny methods. To test this hypothesis, we increased the number of clusters to correspond to the number of serovars (k = 37). If no virulence subpopulations are present (within serovar variance is less than between serovar variance), each of the 37 clusters should contain a majority of one serovar (see methods). However, we found 11 serovars with virulence subpopulations (Table 2). The full list of subpopulation designations is provided in S6 Table. To test if virulence subpopulations may correspond to phenotypic differences in case presentation, we computed the proportion of clinical infections resulting in extraintestinal infections for each serovar subpopulation for sequenced strains with case presentation in the FoodNet surveillance system. Two serovars yielded significant differences in invasiveness between serovar subpopulations. *S.* Infantis split into two subpopulations (subpopulation 18: n = 145, subpopulation 20: n = 243) as shown in Fig 6A. The genome assembly size for subpopulation 18 isolates was significantly longer (4.98 Mb vs. 4.68 Mb, p-value < 2.2E-16, Mann-Whitney U test) (Fig 6B) than isolates from subpopulation 20. Of the 388 *S.* Infantis genome assemblies in the analysis set, 242 had associated clinical presentation data from FoodNet split evenly between the two subpopulations (n = 121, n = 121). Isolates from subpopulation 18 were more than twice as likely to result in extraintestinal clinical infections than isolates from subpopulation 20 (RF: 2.06, 99% CrI: 1.122 – 3.778) (Fig 6C). There was an association between subpopulation 18 isolates and older patients (median age 56.1 years) when compared to subpopulation 20 isolates (median age 36.4 years) (p-value: 5.00E-6, Mann-Whitney U test) (Fig 6D).

**Fig 6.**
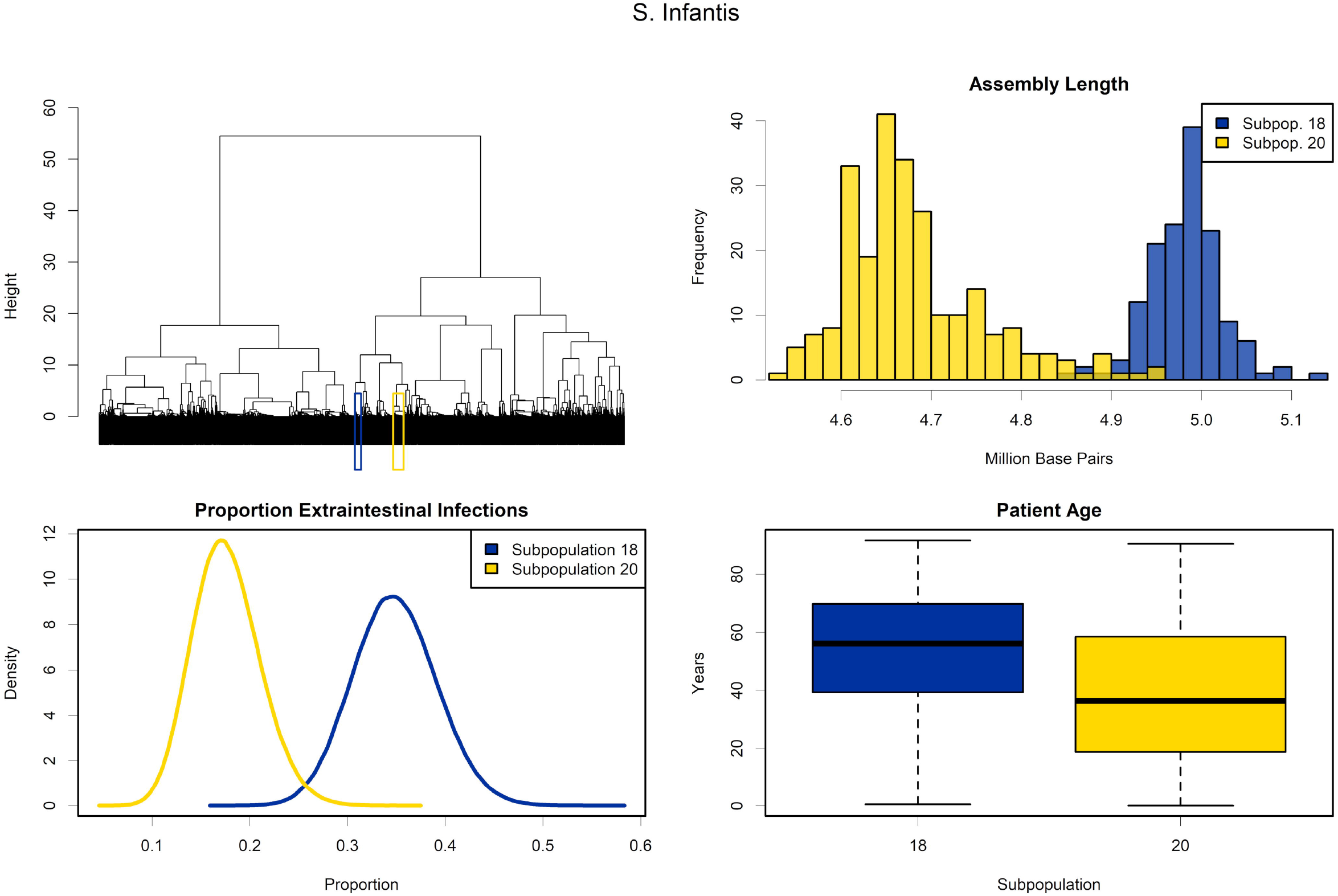
Description of two *S.* Infantis virulence subpopulations. (A) Dendrogram highlighting the locations of the two *S.* Infantis virulence subpopulations within the greater population of 12,337 *S. enterica* isolates. (B) Histograms of the assembly lengths for the respective subpopulations. (C) Proportion of extraintestinal infections among illnesses caused by the two subpopulations (FoodNet data). (D) Boxplots of the distribution of patient age in infections caused by the two subpopulations (FoodNet data).

**Table 2.** Within serovar virulence subpopulations.

| Serovar | Total Serovar Count | Subpopulation A |  |  | Subpopulation B |  |  |
| --- | --- | --- | --- | --- | --- | --- | --- |
|  |  | Subpopulation ID | Genome Count | Proportion of Total Serovar Population | Subpopulation ID | Genome Count | Proportion of Total Serovar Population |
| I 1,4,[5],12:b:- | 83 | 30 | 30 | 0.36 | 29 | 52 | 0.63 |
| I 1,4,[5],12:i:- | 530 | 9 | 294 | 0.55 | 2 | 163 | 0.31 |
| Infantis | 388 | 20 | 243 | 0.63 | 18 | 145 | 0.37 |
| Kentucky | 169 | 32 | 89 | 0.53 | 21 | 80 | 0.47 |
| Montevideo | 1341 | 14 | 503 | 0.38 | 11 | 838 | 0.62 |
| Muenchen | 392 | 12 | 133 | 0.34 | 6 | 195 | 0.50 |
| Newport | 1751 | 6 | 427 | 0.24 | 1 | 926 | 0.53 |
| Oranienburg | 137 | 14 | 32 | 0.23 | 11 | 103 | 0.75 |
| Reading | 60 | 37 | 28 | 0.47 | 29 | 28 | 0.47 |
| Saintpaul | 202 | 29 | 53 | 0.26 | 27 | 134 | 0.66 |
| Typhimurium | 1106 | 16 | 477 | 0.43 | 2 | 460 | 0.42 |

We hypothesized that the approximately 300kb difference between the assembly lengths of the *S.* Infantis subpopulations may be due to the presence of the pESI plasmid previously identified in *S*. Infantis(36). After checking all isolates for the presence of this plasmid,144 out of 145 *S.* Infantis isolates annotated to subpopulation 18 and 0 out of 243 isolates from subpopulation 20 were putatively positive for pESI plasmids. Only one isolate, a *S.* Muenchen, was putatively positive for the pESI plasmid outside of the *S.* Infantis 18 subpopulation.

Two subpopulations represented approximately 85% of the total *S.* Typhimurium population in the analysis set, which we analyzed further (Fig 7A). Similar to the *S.* Infantis subpopulations, the two subpopulations yielded significantly different genome assembly lengths (subpopulation 2: 4.90 Mb, subpopulation 16: 4.85 Mb, p-value < 2.2E-16, Mann-Whitney U test) (Fig 7B). However, the assembly difference of approximately 5kb between the *S.* Typhimurium subpopulations is far less dramatic than the approximately 300kb difference observed between *S.* Infantis subpopulations. 668 of the 937 *S.* Typhimurium isolates in subpopulations 2 (n = 359) and 16 (n = 309) have clinical case presentation data. Subpopulation 2 isolates presented as double the extraintestinal infections than subpopulation 16 isolates (RF 2.11, 99% CrI: 1.109 – 4.016) (Fig 7C). In contrast with the *S.* Infantis subpopulations, the age of patients was not significantly different between the two subpopulations (p-value 0.97, Mann-Whitney U test) (Fig 7D).

**Fig 7.**
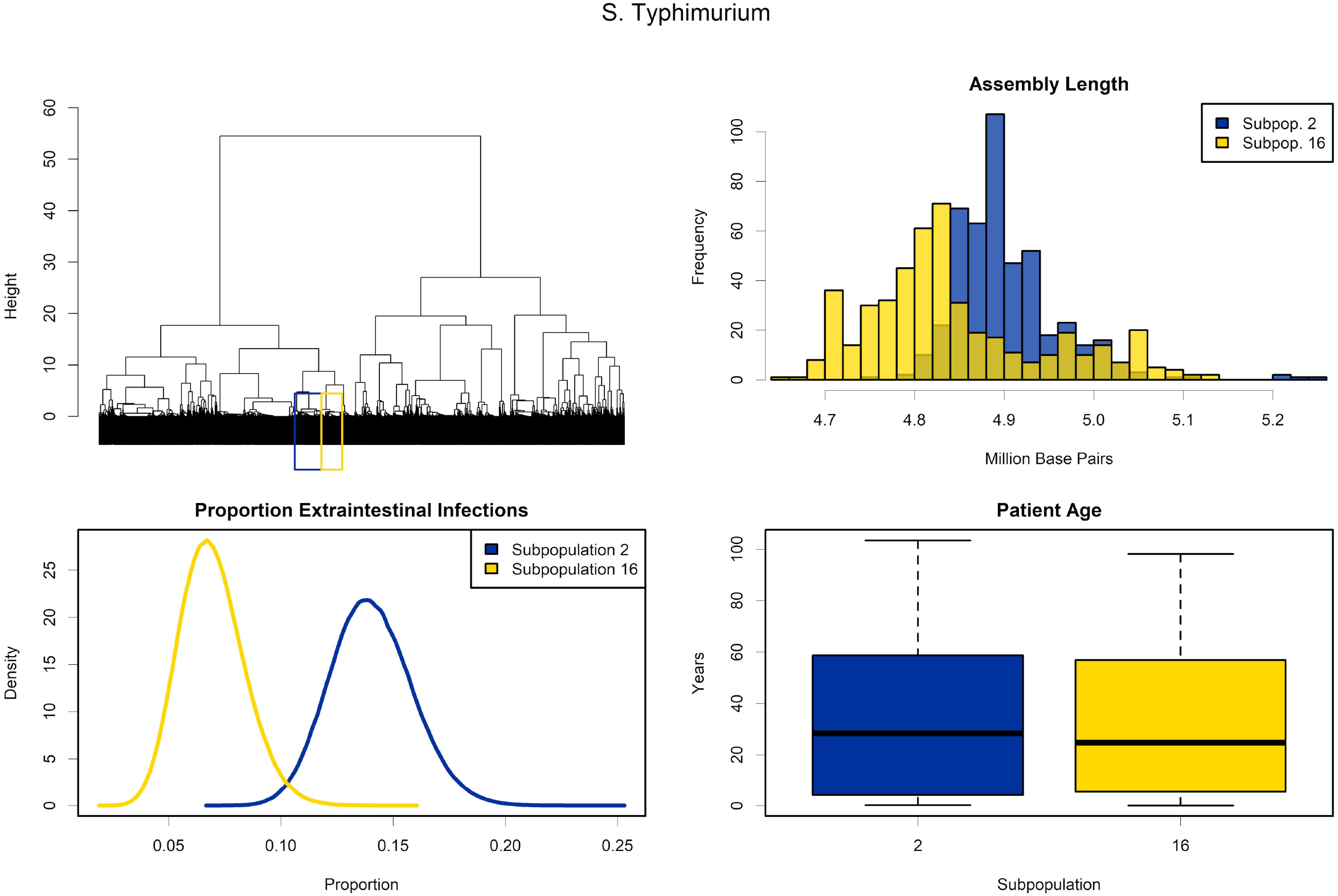
Description of two *S.* Typhimurium virulence subpopulations. (A) Dendrogram highlighting the locations of the two *S.* Typhimurium virulence subpopulations within the greater population of 12,337 *S. enterica* isolates. (B) Histograms of the assembly lengths for the respective subpopulations. (C) Proportion of extraintestinal infections among illnesses caused by the two subpopulations (FoodNet data). (D) Boxplots of the distribution of patient age in infections caused by the two subpopulations (FoodNet data).

Table 2 legend: Serovars found to contain at least two subpopulations, each representing greater than 0.20 of the total serovar population. Subpopulations were identified by increasing the number of clusters to match the number of serovars (k = 37). Provided is the subpopulation ID’s, the number of genomes resident within each subcluster, and the proportion of the total population the subcluster represents. Note, that the subpopulations may not represent the total combined population of the serovar in the analysis set.

## Discussion

The pathogenesis of *S. enterica* is only partially understood, and how different serovars generate distinct disease pathologies is also not well-defined. To better understand how serovars group together based on virulence factor gene carriage, we describe a novel methodology that allowed for rapid identification of serovars of public health concern. Compared to methods used in previous studies [23,24], this scalable genomic approach allowed us to generate a measure of relatedness for a large number of *S. enterica* isolates in a computationally efficient manner and group them using established hierarchical clustering methods [41]. While we considered other clustering methods such as logistic principal component analysis and k-means clustering, we chose the unsupervised random forest approach because it is more robust to outliers, non-parametric, and aggregates results from many models rather than basing inference on a single, “best” model. Our method cannot be read as a traditional phylogeny of evolutionary process but rather as a snapshot of the current virulence potential of more than 12,000 isolates retrieved from humans, bovine animals, and beef products. We did not employ a traditional phylogeny because we were chiefly interested in the current state of potential virulence that consumers are exposed to through beef products, rather than in the evolutionary development of such virulence. As such, we wanted to capture the largest possible number of isolates in a computationally-efficient manner.

We contend that this method is pertinent to virulence loci found with SPI as the regions are subject to horizontal gene transfer [45,46]. Common methods to differentiate serovars typically rely on the alignment of core genes or single nucleotide polymorphisms (SNP) identified against reference assemblies [6,24,47,48]. These methods must rely on *post hoc* analysis to determine if two evolutionary similar strains have acquired virulence factor genes which may correspond to differences in case presentation as witnessed in *S.* Infantis and *S.* Typhimurium. Demarcating isolates by the presence/absence of virulence factors identified a cluster of higher virulence serovars that accounts for a large proportion of sporadic cases, beef-associated, and total foodborne outbreaks compared to the lower virulence cluster. The higher occurrence of beef-associated outbreaks occurs despite a much lower frequency of isolation from regulatory beef samples relative to serovars from the other clusters combined. Our method, in combination with quantitative risk assessment techniques could be used to account for the relative exposure to serovars (e.g., via different food consumption) and the resultant probability of disease.

FoodNet isolates are the basis of the salmonellosis incidence calculation, and the dataset does not provide source attribution. Therefore, serovars from the sporadic human clinical data likely result from multiple exposure sources (poultry, beef, vegetables, etc.). Despite this potentially diverse source of *S. enterica* isolates, most serovars resided in one of the five major clusters (including beef and bovine isolates) suggesting that basal virulence factor gene carriage is conserved within serovars across sources.

Interestingly, our method identified a group of virulence factors involved in attachment to host cells or outer membrane structure as the most differential genes between virulence groups (Fig 5). Further, many of these adhesion operons originated from Enterobacteriaceae other than *Salmonella*. Use of only putative *Salmonella* virulence factors from PATRIC [32] and the Virulence Factor Database [31] would not have annotated the open reading frames highlighting the need for expanding the putative virulence factors of *S. enterica* outside of the genera to members of Enterobacteriaceae. In contrast, we found two differentiating operons, *lpf* (long polar fimbriae) and *rfb,* from different virulence categories in significantly higher proportions in cluster 1. The higher proportion of *lpf* genes, which fall into the adhesion virulence category, is notable as the operon has been associated with *S. enterica* binding the Peyer’s patches, namely the M-cells found within the lymphoid organs [49,50]. This may potentiate the higher infectivity of cluster 1 serovars as recent work with the Type Three Secretion systems (T3SS) of *S. enterica* (involved with the introduction of effector proteins to the cytoplasm of host cells) suggest that the structure does not penetrate the cytoplasmic membrane like a syringe, but requires tension and adopts a “tent-pole” like structure [51]. If tension is required for the function of the T3SS, enhanced binding to M-cells mediated by the *lpf* operon may be one reason cluster 1 has a higher incidence rate of domestically acquired sporadic cases. The *lpf* operon has not only been implicated in *S. enterica* infections; strains of *Escherichia coli* O157:H7 with mutations in the *lpf* operon show decreased attachment and colonization in both in vitro [52] and in vivo [53] models again show the interplay of virulence mechanisms between Enterobacteriaceae. The roles of *rfb* genes, which were primarily classified in the survival/host persistence virulence category, are not as well investigated as the *lpf* operon but appear to be involved in the biosynthesis of the O-antigen and lipopolysaccharide structuring. A recent report suggests that the full complement of *rfb* genes leads to higher virulence in experimentally infected chickens [54].

Examining virulence gene catalogues not only identified large, serovar level clusters but also, by altering the cluster number (k value), virulence subpopulations within serovars. With the current method, it cannot be ascertained whether the virulence subpopulations represent polyphyletic clades within serovars as it cannot be interpretated as a phylogeny. However, by applying a top-down approach, the presence of increased virulence capacity can be readily identified. The two subpopulations of *S.* Infantis present over a two-fold difference in probability of extraintestinal infections. *S.* Infantis has been rapidly increasing in incidence in Israel and previous studies suggest that the addition of a virulence megaplasmid pESI could be responsible [55]. The mean difference between the two subpopulations was approximately 300kb, similar in length to the pESI plasmid (280 kb), and querying *S.* Infantis isolates against a database of marker genes revealed that isolates in the subpopulation with longer assemblies are putatively positive for pESI presence. In addition, the pESI plasmid carries genes necessary for the synthesis of yersiniabactin [55], a siderophore dependent iron uptake system commonly observed in *Yersinia pestis*. The eight genes comprising the *ybt* operon are resident in every strain of the higher invasive cluster of *S*. Infantis, and only two out of 243 isolates from the lower invasive cluster contain the operon. Iron is an essential nutrient for *S. enterica* replication during systemic infections [56]. A previous study suggests that co-infections of Malaria and *S. enterica* leads to more systemic infections as excess iron is released upon the lysis of red blood cells, liberating the metal for use by *S. enterica* [57]. Increased iron availability, due to the addition of yersiniabactin, may be one factor for the almost double rate of extraintestinal infections of *S.* Infantis cluster 18 compared to *S.* Infantis infections without this plasmid.

The methods employed here cannot identify virulence changes due to sequence variations within virulence loci. Variants of the *macA* and *macB* genes in African strains of *S.* Typhimurium sequence-type 313 may have higher invasiveness in human patients and increased survival against challenge with antimicrobial peptides [58]. Others have identified virulence gene alleles that may correspond to pathogenicity differences [59]. The method employed identifies virulence genes against a non-redundant database using BLASTP, so alleles with variation less than 10% sequence identity will be collapsed into the same gene annotation. Furthermore, we did not consider pseudogene formation of virulence genes. Previous work suggests that pseudogenes in *S. enterica* genomes do not follow neutral evolution (random genetic drift, as in many Eukaryotes) but are readily lost from the chromosome [60]. However, pseudogene formation of the *sseI/srfH* secreted effector protein leads to hyperdissementation of ST313 *S.* Typhimurium in experimentally infected mice [61]. The role of pseudogene formation and the pathogenesis needs more study, and the addition of pseudogene information could further improve virulence classifications. Additionally, we chose to focus our analysis on human, bovine, and beef isolates from the US. It is probable given the diversity of *S. enterica* that all virulence patterns and serovar subpopulations are not represented in this work.

*S. enterica* is a diverse pathogen. Yet, most risk assessments and food safety regulations informed by these assessments only separate Typhoidal Salmonellosis and non-Typhoidal Salmonellosis, treating serovars as a homogenous unit [62-64]. Our results suggest that strains with the highest incidence of domestically acquired sporadic cases and outbreaks of human infections share a common virulence repertoire. Control and surveillance programs devoting more resources to clinically relevant serovars might result in increased public health gains, but such interventions must be evaluated using quantitative risk assessment methods.

Furthermore, serovar virulence cannot be considered homogenous in all cases as observed with *S.* Infantis and *S.* Typhimurium. Although attributing virulence to specific genes was beyond the scope of this study, our analysis could inform further research to identify *Salmonella* genes associated with severe illness.

## Data Availability

All data produced in the present study will be available in a public repository upon acceptance of this manuscript to a peer-reviewed journal.

## Acknowledgements

G.F, J.P., D.T., S.C., and F.Z. are employed by EpiX Analytics. R.P. is a Senior Scientific Advisor for EpiX Analytics. This work utilized the RMACC Summit supercomputer, which is supported by the National Science Foundation (awards ACI-1532235 and ACI-1532236), the University of Colorado Boulder, and Colorado State University. The Summit supercomputer is a joint effort of the University of Colorado Boulder and Colorado State University. FoodNet Data: The findings and conclusions in this report are those of the author(s) and do not necessarily represent the official position of the Centers for Disease Control and Prevention.

## Supporting information

**S1 Fig. Addition of a sixth virulence cluster.** (A) Dendrogram depicting the hierarchical relationship between 12,337 S. enterica genome assemblies based upon virulence factor gene carriage with six virulence clusters superimposed on top. (B) Heatmap of serovar proportion within each of the six respective virulence clusters. Rows are clustered using Ward’s method. (C) Characteristics of the six virulence clusters: cluster stability - Jaccard similarity of 10,000 non-parametric bootstraps, Number of Genomes - depicting the number of *S*. *enterica* genomes constituent in each cluster, and number of serovars (within cluster serovar proportion > 0.5) in each cluster.

**S1 Table. Metadata for the analysis set of genomes and SISTR serovar prediction**. Metadata for the contig assemblies used in the analysis including results of the in silico serovar prediction for the analysis set genomes from the SISTR software.

**S2 Table. Serovar virulence cluster designations.** Virulence cluster designations (k = 5) for the 37 serovars in the analysis set.

**S3 Table. Full list of putative virulence loci considered in the random forest model.** Gene name, locus tag, database source, Genus origin, gene product and classification for the 182 putative virulence factor loci used in the random forest model.

**S4 Table. Epidemiological indicators computed for each virulence cluster**. Estimates of: incidence of domestically acquired sporadic cases per 100k people per year, hospitalization proportion given infection, proportion positive in FSIS testing of US beef products (MT43, MT60, MT64), extraintestinal proportion given infection, and mortality proportion given infection.

**S5 Table. Virulence factors present in all isolates**. List of virulence factor genes (n=68) present in all isolates.

**S6 Table. Isolate virulence subpopulation cluster designations.** Subpopulation cluster designations (k = 37) for the 12,337 contig assemblies in the analysis set.

